# SARS-CoV-2 specific antibody responses in COVID-19 patients

**DOI:** 10.1101/2020.03.18.20038059

**Authors:** Nisreen M.A. Okba, Marcel A. Müller, Wentao Li, Chunyan Wang, Corine H. GeurtsvanKessel, Victor M. Corman, Mart M. Lamers, Reina S. Sikkema, Erwin de Bruin, Felicity D. Chandler, Yazdan Yazdanpanah, Quentin Le Hingrat, Diane Descamps, Nadhira Houhou-Fidouh, Chantal B. E. M. Reusken, Berend-Jan Bosch, Christian Drosten, Marion P.G. Koopmans, Bart L. Haagmans

## Abstract

A new coronavirus, SARS-CoV-2, has recently emerged to cause a human pandemic. Whereas molecular diagnostic tests were rapidly developed, serologic assays are still lacking, yet urgently needed. Validated serologic assays are important for contact tracing, identifying the viral reservoir and epidemiological studies. Here, we developed serological assays for the detection of SARS-CoV-2 neutralizing, spike- and nucleocapsid-specific antibodies. Using serum samples from patients with PCR-confirmed infections of SARS-CoV-2, other coronaviruses, or other respiratory pathogenic infections, we validated and tested various antigens in different in-house and commercial ELISAs. We demonstrate that most PCR-confirmed SARS-CoV-2 infected individuals seroconverted, as revealed by sensitive and specific in-house ELISAs. We found that commercial S1 IgG or IgA ELISAs were of lower specificity while sensitivity varied between the two, with IgA showing higher sensitivity. Overall, the validated assays described here can be instrumental for the detection of SARS-CoV-2-specific antibodies for diagnostic, seroepidemiological and vaccine evaluation studies.

## Introduction

In December 2019, a new coronavirus (CoV) emerged in China to cause an acute respiratory disease known as coronavirus disease 19 (COVID-19) (1). The virus was identified to be a betacoronavirus related to severe acute respiratory syndrome coronavirus (SARS-CoV) and thus, was named SARS-CoV-2 (2). In less than two decades, this virus is the third known coronavirus to cross the species barrier and cause severe respiratory infections in humans following SARS-CoV in 2003 and Middle East respiratory syndrome in 2012, yet with unprecedented spread compared to the earlier two. Due to the rapid rise in number of cases and uncontrolled and vast worldwide spread, the WHO has declared SARS-CoV-2 a pandemic. As of March 14th 2020, the virus has infected over 130,000 individuals in 122 countries, 3.7% of which had a fatal outcome (3). The rapid identification of the etiology and the sharing of the genetic sequence of the virus, followed by international collaborative efforts initiated due to the emergence of SARS-CoV-2 have led to the rapid availability of real-time PCR diagnostic assays that support the case ascertainment and tracking of the outbreak (4). The availability of these has helped in patient detection and efforts to contain the virus. However, specific and validated serologic assays are still lacking at the moment and are urgently needed to understand the epidemiology of SARS-CoV-2.

Validated serologic assays are crucial for patient contact tracing, identifying the viral reservoir hosts and for epidemiological studies. Epidemiological studies are urgently needed to help uncover the burden of disease, in particular, the rate of asymptomatic infections, and to get better estimates on morbidity and mortality. Additionally, these epidemiological studies can help reveal the extent of virus spread in households, communities and specific settings; which could help guide control measures Serological assays are also needed for evaluation of the results of vaccine trials and development of therapeutic antibodies. Among the four coronavirus structural proteins, the spike (S) and the nucleocapsid (N) are the main immunogens (5). Here, we describe development of serological assays for the detection of virus neutralizing antibodies and antibodies to the nucleocapsid (N) protein and various spike (S) domains including the S1 subunit, and receptor binding domain (RBD) of SARS-CoV-2 in ELISA format. Using a well-characterized cohort of serum samples from PCR-confirmed SARS-CoV-2 and patients PCR-confirmed to be infected with seasonal coronaviruses and other respiratory pathogens, we validated and tested various antigens in different platforms developed in-house as well as a commercial platform.

## Materials and methods

### Serum Samples

#### EMC samples

Serum samples were collected from PCR-confirmed mild and severe COVID-19 patients (**Table 1**) from France in accordance with the local ethical approvals. Samples used for assay validation were from persons PCR-diagnosed infections with of human coronaviruses (HCoV-229E, NL63 or OC43), SARS, MERS, or with a range of other respiratory viruses (**Table 1**) as published previously (6). Samples from patients with recent CMV, EBV or mycoplasma pneumoniae infection were included as these have a higher likelihood of causing false positive results. We used serum samples from 45 healthy blood donors (Cohort A) as negative controls; Sanquin Blood Bank (Rotterdam, the Netherlands) obtained written informed consent for research use. All samples were stored at -20°C until use. The use of serum samples from the Netherlands was approved by the local medical ethical committee (MEC approval: 2014–414). Serum samples from SARS patients (7) were kindly provided by professor Malik Peiris, Hong Kong University.

**Table 1.**
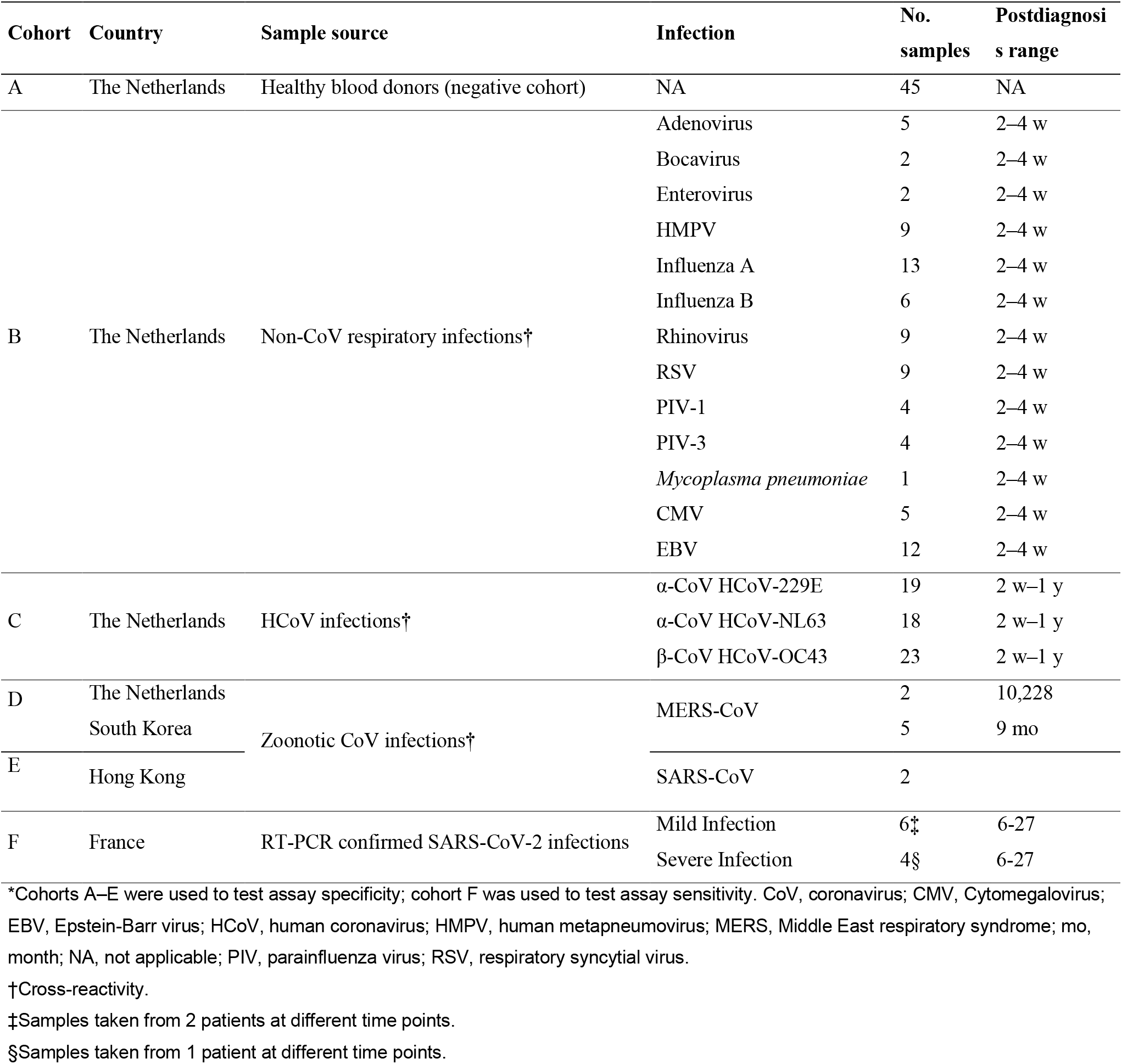
Cohorts used to validate the specificity and sensitivity of assays for SARS-CoV-2

#### Berlin samples

All serum samples from COVID-19 PCR-confirmed cases (*n* = 9) were previously analyzed by recombinant SARS-CoV-2 Spike protein-based immunofluorescence test and plaque reduction neutralization (Wölfel et al submitted manuscript, doi:https://doi.org/10.1101/2020.03.05.20030502). Sera were tested as part of an extended diagnostic regimen upon informed written consent of patients. All non-SARS-CoV-2 sera (*n*=31) stemmed from the serum collection of the national consiliary laboratory for coronavirus detection at Charité, Berlin, Germany and were provided for diagnostic proposes upon informed written consent. The collection contained follow-up sera from PCR-confirmed human cases: *n*=4 HCoV-229E, *n*=3 HCoV-HKU1, *n*=7 HCoV-OC43, *n*=3 MERS-CoV, *n*=6 HCoV-NL63, *n*=3 SARS-CoV and *n*=6 common cold CoV antibody positive sera.

### Protein expression

The spike ectodomains of SARS-CoV-2 (residues 1–1,213; strain Wuhan-Hu-1; GenBank: QHD43416.1), SARS-CoV (residues 1–1,182; strain CUHK-W1; GenBank: AAP13567.1) and MERS-CoV (residues 1-1262; strain EMC; GenBank: YP_009047204.1) were expressed in HEK-293T cells with a C-terminal trimerization motif and Strep-tag using the pCAGGS expression plasmid. Likewise, the SARS-CoV-2 S1 subunit or its subdomains (S;S1, residues 1-682; S1^A^, residues 1-294; RBD, residues 329-538; GenBank: QHD43416.1) were expressed in 293T cells as described in (Chunyan Wang et al., submitted manuscript; doi: https://doi.org/10.1101/2020.03.11.987958). Spike S1 proteins of other HCoVs, –HKU1 (residues 1–750), –OC43 (residues 1–760), NL63 (residues 1–717), 229E (residues 1–537), SARS-CoV (residues 1–676), and MERS-CoV were expressed as described earlier (6, 8). All the recombinant proteins were affinity purified from the culture supernatant by protein-A sepharose beads (Cat.no: 17-0780-01, GE Healthcare) or streptactin beads (Cat.no: 2-1201-010, IBA) purification. Purity and integrity of all purified recombinant proteins was checked by coomassie stained SDS-PAGE.

### PRNT

PRNT was used as a reference for this study, because neutralization assays are the standard for CoV serology. We tested serum samples for their neutralization capacity against SARS-CoV-2 (German isolate; GISAID ID EPI_ISL 406862; European Virus Archive Global # 026V-03883) by plaque-reduction neutralization test (PRNT) as previously described for with some modifications (9). Heat-inactivated samples were 2-fold serially diluted in DMEM medium supplemented with NaHCO_3_, HEPES buffer, penicillin, streptomycin, and 1% fetal bovine serum, starting at a dilution of 1:10 in 50 μL. Fifty μL of the virus suspension (400 spot forming units) was added to each well and incubated at 37°C for 1 h. Following incubation, the mixtures were added on Vero-E6 cells and incubated at 37°C for 1 more hour. The cells were then washed and further incubated in medium for 8 h. After the incubation, the cells were fixed and stained with polyclonal rabbit anti-SARS-CoV antibody (Sino Biological). The cells were then fixed and stained using a rabbit anti-SARS-CoV serum and a secondary peroxidase-labelled goat anti-rabbit IgG (Dako). The signal was developed using a precipitate forming TMB substrate (True Blue, KPL) and the number of infected cells per well were counted using the ImmunoSpot® Image analyzer (CTL Europe GmbH). The serum neutralization titer is the reciprocal of the highest dilution resulting in an infection reduction of >50% (PRNT_50_). A titer of >20 was considered to be positive.

The PRNT for the German sera was done as described previously using Vero E6 cells (Wölfel et al submitted manuscript, doi:https://doi.org/10.1101/2020.03.05.20030502) (10). Prior to PRNT patient sera were heat-inactivated at 56°C for 30 minutes. For each dilution step (duplicate), patient sera were diluted in 200 ul OptiPro and mixed 1:1 with 200 ul virus solution containing 100 plaque forming units. The 400 ul serum-virus solution was vortexed gently and incubated at 37°C for 1 hour. Each 24-well was incubated with 200 ul serum-virus solution. After 1 hour at 37°C the supernatants were discarded, the cells were washed once with PBS and supplemented with 1.2% Avicel solution in DMEM. After 3 days at plates were fixed and inactivated using a 6% formaldehyde/PBS solution and stained with crystal violet.

### ELISA

We performed Anti-SARS-CoV-2 IgG and IgA ELISA using beta-versions of two commercial kits (EUROIMMUN Medizinische Labordiagnostika AG, https://www.euroimmun.com) and performed the assay according to manufacturer’s protocol. Reagent wells of both assays are coated with recombinant structural protein (S1 domain) of SARS-CoV-2. The optical density (OD) was detected at 450 nm, and a ratio of the reading of each sample to the reading of the calibrator, included in the kit, was calculated for each sample (OD ratio). As the beta-version of the kit awaits CE validation, we determined an in-house cut-off value based on the mean background reactivity of all SARS-CoV-2-negative sera in the study multiplied by 3. In case of IgA this was OD ratio = 0.9 and for IgG OD ratio = 0.3.

We performed the inhouse ELISAs by coating 96-well microtiter ELISA plates with in-house produced spike antigens (S or S1 of SARS-CoV-2, SARS-CoV or MERS-CoV; or SARS-CoV-2 S1^A^, or RBD proteins) or SARS-N (Sinobiological) in PBS overnight at 4°C. Following blocking, diluted serum (1:100 or 2-fold serially diluted for titers) was added and incubated at 37°C for 1h. Antigen-specific antibodies were detected using peroxidase-labeled rabbit anti– human IgG (Dako, https://www.agilent.com) and TMB as a substrate. The absorbance of each sample was measured at 450 nm. We set cutoffs at 6 standard deviations above the mean value of the negative cohort.

### S1 Protein Microarray

Serum samples were previously tested for antibodies against the S1 of different coronaviruses as described earlier (6).

### Statistical Analysis

The correlations between antibody responses detected by different ELISAs were and those detected by PRNT, as the gold standard for CoV serology, were analyzed using GraphPad Prism version 8 (https://www.graphpad.com).

## Results

We evaluated SARS-CoV-2 specific antibody responses in severe and mild cases using serum samples collected at different times post-disease onset from three French PCR-confirmed CoVID-19 patients. We tested sera for SARS-CoV-2 specific antibodies using different ELISAs. Following infections, all three patients seroconverted between days 13 and 21 post onset of disease (**Figure 1**), and antibodies were elicited against the SARS-CoV-2 S and S1 subunit including the N-terminal (S1^A^) domain and the receptor binding domain (RBD). Since the N protein of SARS-CoV-2 is 90% similar to that of SARS-CoV (**Table 2**), we used SARS-CoV N as an antigen to test for SARS-CoV-2 N-directed antibodies in an ELISA format. We found that, following infection, antibodies were elicited against the N protein and when tested in a PRNT assay these sera were able to neutralize SARS-CoV-2. We observed cross-reactivity with the SARS-CoV S and S1 proteins, and to a lower extent with MERS-CoV S protein, but not with the MERS-CoV S1 protein (**Figure 1 G-H**). This was evident from analyzing the degree of similarity of the different CoV S protein domains to their corresponding SARS-CoV-2 proteins (**Table 2**), where SARS-CoV showed high similarities in all different S domains. The analysis showed that the spike S2 subunit is more conserved among CoVs and thus plays a role in the cross-reactivity seen when the whole S was used as an antigen. Thus, S1 is a more specific than S as an antigen for SARS-CoV-2 serological diagnosis.

**Table 2.** Percentage amino acid identity of coronavirus spike and nucleocapsid proteins to SARS-CoV-2 proteins.

|  |  | *N* | *S* | *S1* | *S2* | *S1<sup>A</sup>* | *RBD* |
| --- | --- | --- | --- | --- | --- | --- | --- |
| *Beta-CoV* | SARS-CoV | 90 | 77 | 66 | 90 | 52 | 73 |
|  | MERS-CoV | 49 | 33 | 24 | 43 | nd | nd |
|  | HCoV-OC43 | 34 | 33 | 25 | 42 | nd | nd |
|  | HCoV-HKU1 | 34 | 32 | 25 | 40 | nd | nd |
| *Alpha-CoV* | HCoV-229E | 28 | 30 | 24 | 35 | nd | nd |
|  | HCoV-NL63 | 29 | 28 | 21 | 36 | nd | nd |

SARS-CoV-2, HCoV-OC43, MERS-CoV, HCoV-HKU1, HCoV-NL63, SARS-CoV, HCoV-229E (Acc. nos.
NC\_045512.2, NC\_006213.1, NC\_019843.3, NC\_006577.2, NC\_005831.2, NC\_004718.3, NC\_002645.1).
Protein sequences were aligned using ClustalW. N, nucleocapsid; S, spike; S1, The N-terminal subunit of the spike protein; S2, the C-terminal subunit of the spike protein; S1A, domain A of the spike S1 subunit; RBD, receptor binding domain; nd, not done.

**Figure 1:**
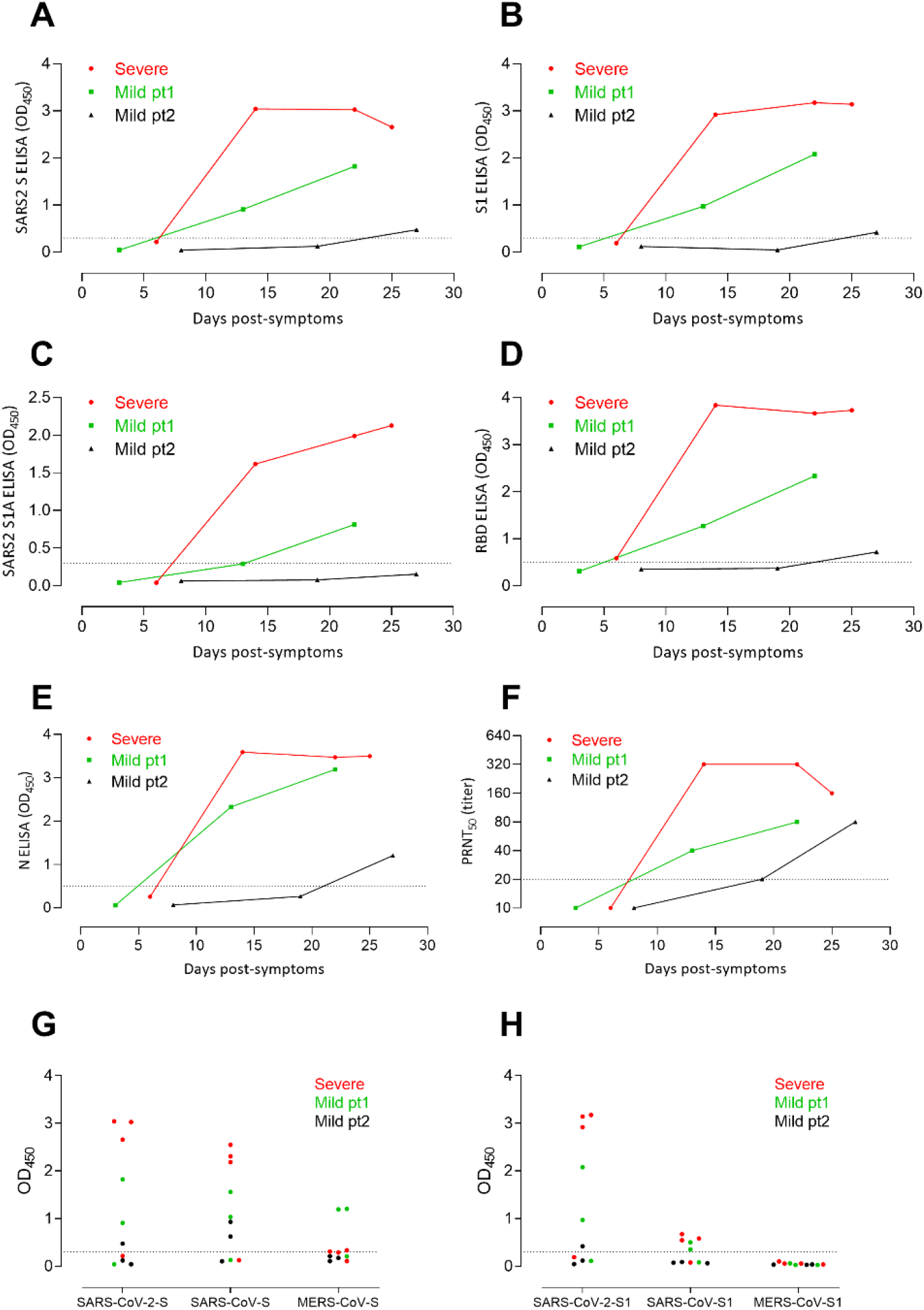
Kinetics of antibody responses against SARS-CoV-2 following infection. We tested one severe (red) and two mild (green and black) SARS-CoV-2 patients for antibody responses against the (A) Spike (S), (B) spike S1 subunit, (C) spike N-terminal (S1^A^) domain, (D) receptor binding domain, (E) nucleocapsid proteins using ELISAs. (F) Virus neutralizing antibodies were tested using the plaque reduction neutralization assay (PRNT). Reactivities of sera form the three patients at different time points against whole spikes (G) and S1 (H) of SARS-CoV-2, SARS-CoV and MERS-CoV were tested by ELISA. SARS-CoV, severe acute respiratory syndrome coronavirus; MERS-CoV, Middle East respiratory syndrome coronavirus;

We further assessed the specificity of the S1 assay using cohorts A-E (**Table 1**) comprising of sera from healthy blood donors (A), PCR-confirmed acute respiratory non-CoV infections (B), acute to convalescent PCR-confirmed alpha- and beta-HCoV infections (C), and PCR-confirmed MERS-CoV (D) and SARS-CoV (E) infections. None of the sera from specificity cohorts A-D were reactive in our in-house S1 ELISA at the set cut-off indicating 100% specificity, whereas sera from SARS-CoV patients cross-reacted (**Figure 2A**). Additionally, the specificity of S1 as an antigen for SARS-CoV-2 serology was further supported by the fact that 87-100 % of the cohort A-C sera included in this study were seropositive for the endemic HCoVs (HCoV-HKU1, HCoV-OC43, HCoV-NL63, and -229E) as determined by the S1 protein microarray (**Figure 2B**). Nonetheless, all were seronegative for SARS-CoV and MERS-CoV. Using the same cohort, we also validated the specificity of the anti-nucleocapsid and anti-RBD IgG ELISAs for detecting SARS-CoV-2 specific antibodies. At the set cut-off, except of SARS-CoV patient sera, none of the control sera tested positive for anti-RBD nor anti-nucleocapsid antibodies (**Figure 2 C,D**), whereas we detected seroconversion among the three COVID-19 patients. These validated ELISAs for different antigens can be useful for epidemiological studies as well as for evaluation of vaccine-induced immune responses.

**Figure 2:**
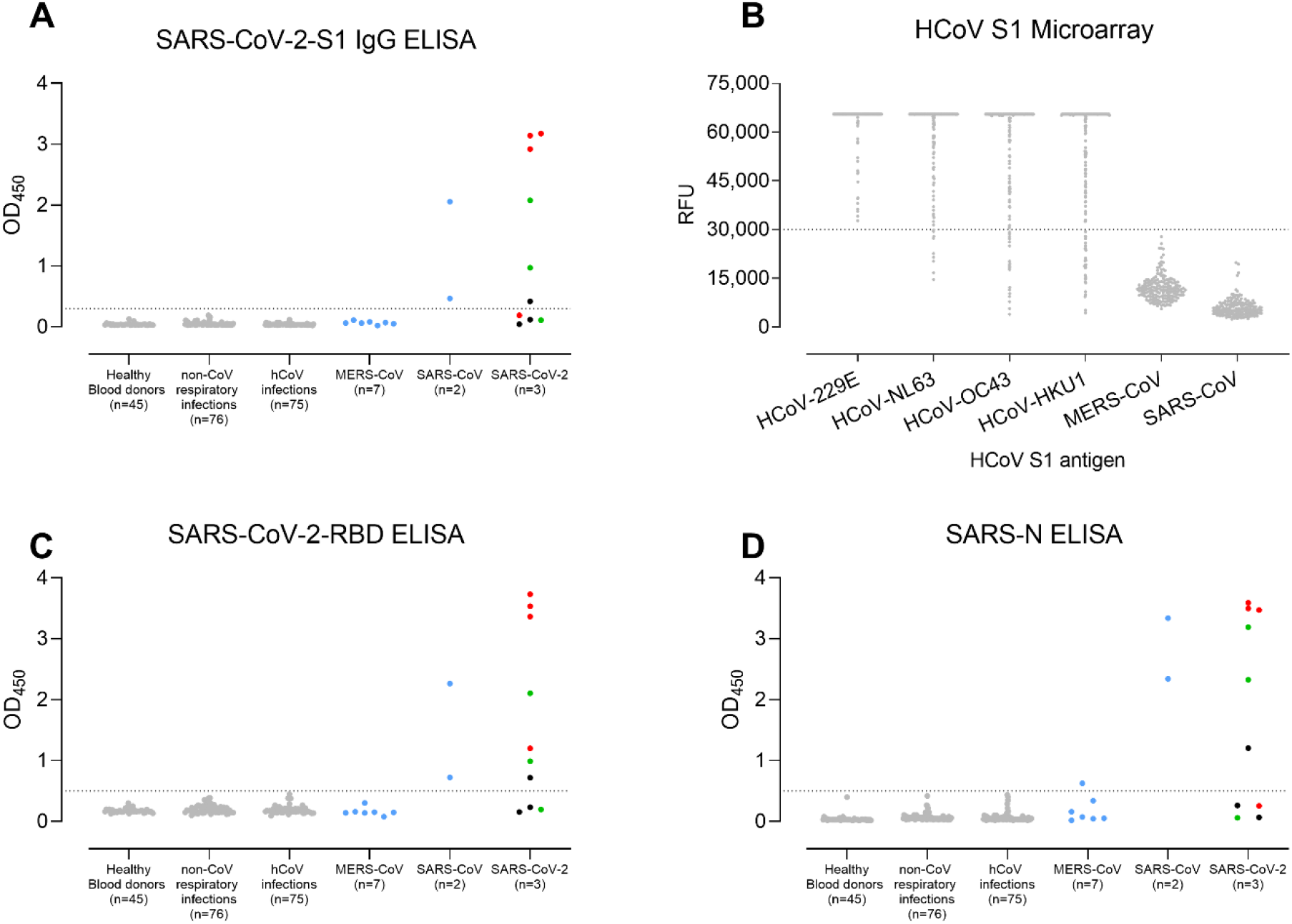
Validation of the use of S1 (A,B), receptor binding domain (RBD; C) and nucleocapsid (N; D)-based ELISAs for detection of SARS-CoV-2 Infections.

Next, we validated the sensitivity and specificity of 2 commercial ELISA kits for detecting S1-specific IgG and IgA antibodies using the same cohort (**Table1, Figure 3**). All three COVID-19 patients had reactive antibodies by both the IgG (6/10 serum samples) and IgA (7/10 serum samples) ELISAs (**Figure 3**). While SARS-CoV patient’s sera were reactive as noted earlier, we also detected reactivity of serum samples from the validation cohorts A-D; 10/203 for IgA and 7/203 for IgG ELISAs. Two HCoV-OC43 (a β-CoV) patients’ sera were reactive in both IgG and IgA ELISA kits. We confirmed the cross-reactivity of the two sera by testing twelve serum samples from both patients collected at different time points, pre- and post-OC43 infection. While all pre-infection sera were negative, all post-infection sera were reactive in both IgG and IgA based ELISAs. We have earlier reported cross-reactivity of these sera in a MERS-CoV S1 IgG ELISA kit (6). Further validation was also done in a different laboratory using 31 serum samples collected from 9 COVID-19 German patients (Wölfel et al submitted manuscript) at different time points (3-23 days post disease onset) as well as a specificity cohort comprising of 18 serum samples from HCoV (4x HCoV-229E, 3x HCoV-HKU1, 4x HCoV-NL63, 7x HCoV-OC43) as well as MERS-CoV (n=3) and SARS-CoV (n=3) infected persons collected 4-56 days post disease onset (**Figure 4**). All 9 COVID-19 patients were previously confirmed to seroconvert at days 6-15 post onset of disease using recombinant immunofluorescence test and PRNT. 8/9 seroconverted patients showed reactivity above the implemented cut-off values in the IgG and IgA ELISA. One patient (**Figure 4**, black line) maintained slightly below the cut-off which might be explained by an overall reduced antibody response of this patient (PRNT_90_=10). Overall, the IgA-based ELISA kit was more sensitive but less specific than the IgG based ELISA kit.

**Figure 3:**
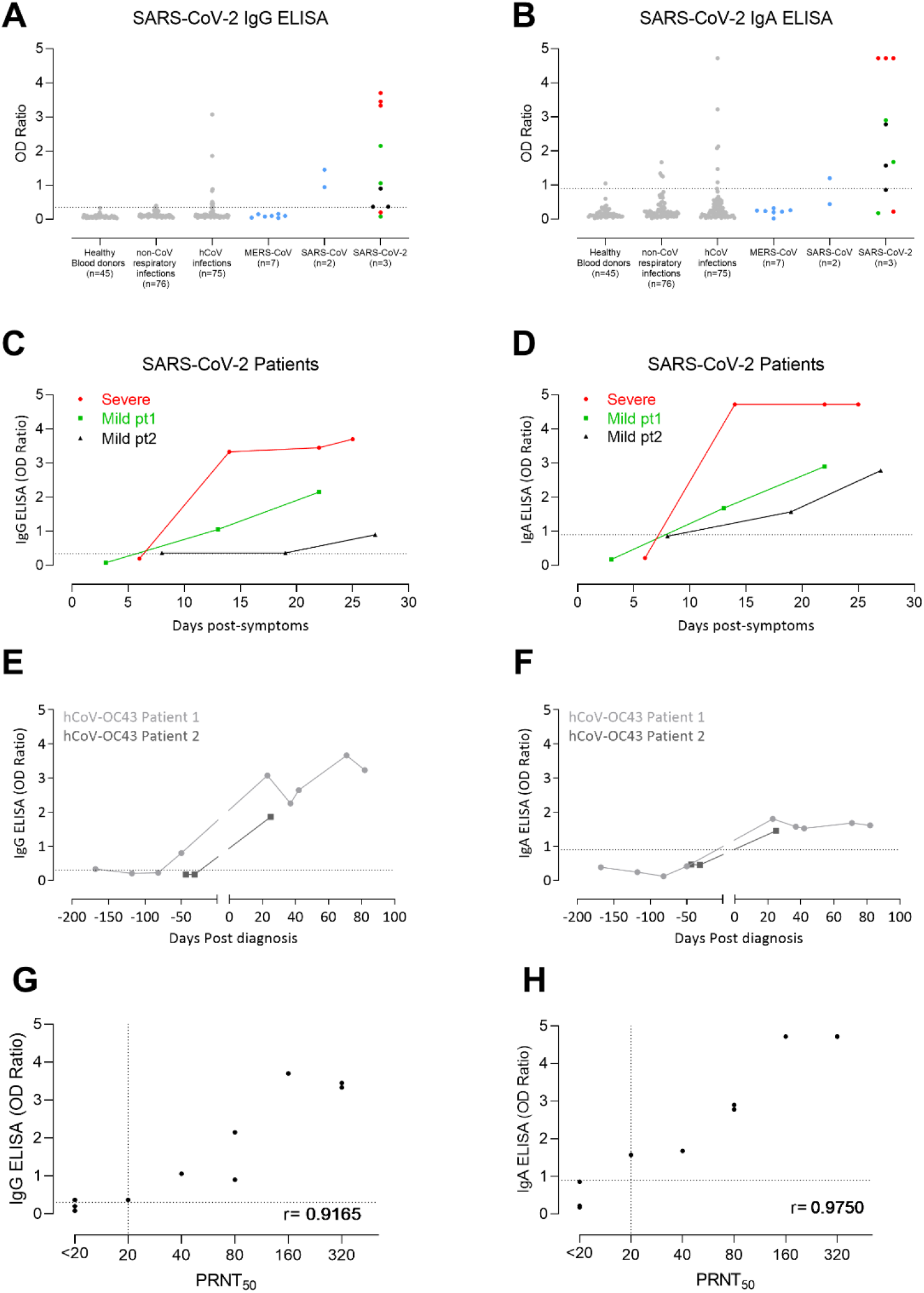
Validation of two commercial ELISAs for the detection of SARS-CoV-2 specific IgG (A,C,E,G) and IgA (B,D,F,H) antibodies. (A,B) validation of the specificity of the two ELISA platforms. Kinetics of antibody responses in three COVID-19 patients (C,D). Cross-reactivity of some HCoV-OC43 sera in the commercial platforms (E,F). Correlation between antibody responses detected by the ELISAs and the plaque reduction neutralization assay (PRNT_50_; G,H).

**Figure 4:**
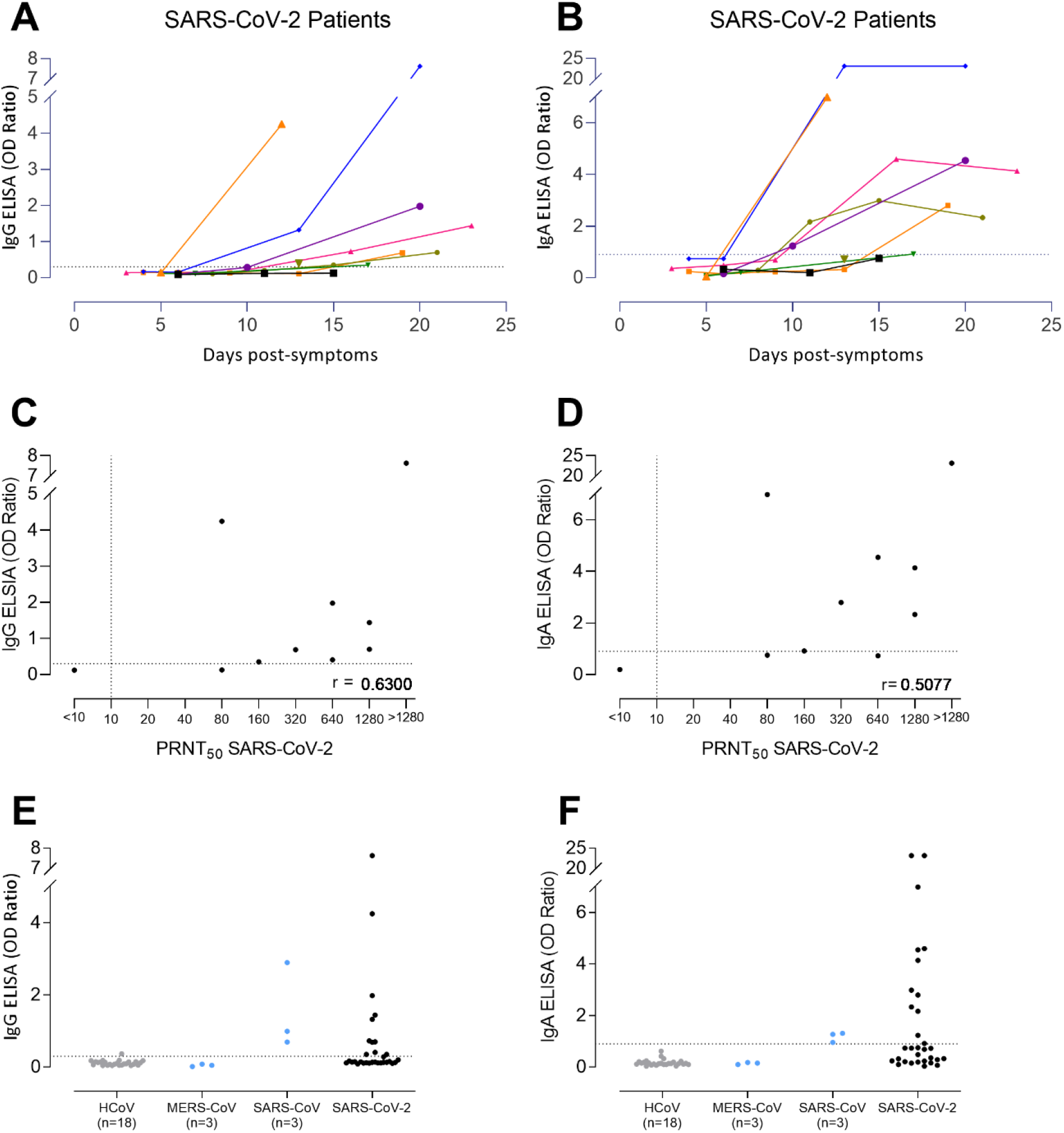
Sensitivity of the two commercial ELISAs for the detection of SARS-CoV-2 specific IgG (A,C,E) and IgA (B,D,F) antibodies. Kinetics of antibody responses in 9 COVID-19 patients from Germany (A,B). Correlation between antibody responses detected by the ELISAs and the plaque reduction neutralization assay (PRNT_50_; C,D). (E,F) The kits were tested for specificity using18 serum samples from HCoV (4x HCoV-229E, 3x HCoV-HKU1, 4x HCoV-NL63, 7x HCoV-OC43) as well as MERS-CoV (n=3) and SARS-CoV (n=3) infected persons.

Finally, we compared the performance of the different ELISAs for the detection of antibodies among PCR-confirmed COVID-19 patients to that of PRNT, as the gold standard for CoV serology (**Table 3**). PRNT_50_ correlated strongly with the different ELISAs, with the commercial IgA showing the strongest correlation followed by the S and N ELISAs indicating their capacity to detect SARS-CoV-2 specific antibodies. However, a larger patient cohort is needed to assess the sensitivity of these platforms.

**Table 3.** Correlations between ODs/OD ratios vs PRNT results of the PCR-confirmed COVID-19 patients tested in Rotterdam (A, n=10 serum samples collected 6-27 days post diagnosis from 3 French COVID-19 patients) and Berlin (B, n=31 serum samples collected 3-23 days post disease onset from 9 German COVID-19 patients).

|  |  |  | Inhouse ELISAs |  |  |  |  | Euroimmun |  |
| --- | --- | --- | --- | --- | --- | --- | --- | --- | --- |
|  |  |  | S1 | N | RBD | S | S1A | IgA | IgG |
| PRNT<sub>50</sub> SARS2 | Spearman r | r | 0.87 | 0.94 | 0.92 | 0.94 | 0.93 | 0.98 | 0.92 |
|  | P value | P (two-tailed) | 0.0021 | 0.0002 | 0.0005 | 0.0002 | 0.0003 | <0.0001 | 0.0005 |
|  |  | P value summary | ** | *** | *** | *** | *** | **** | *** |
| PRNT<sub>90</sub> SARS2 | Spearman r | r | 0.88 | 0.88 | 0.88 | 0.88 | 0.88 | 0.93 | 0.88 |
|  | P value | P (two-tailed) | 0.0024 | 0.0024 | 0.0024 | 0.0024 | 0.0024 | 0.0008 | 0.002 |
|  |  | P value summary | ** | ** | ** | ** | ** | *** | ** |

B
|  |  |  | Euroimmun |  |
| --- | --- | --- | --- | --- |
|  |  |  | IgA | IgG |
| PRNT<sub>50</sub> SARS2 | Spearman r | r | 0.63 | 0.5077 |
|  | P value | P (two-tailed) | 0.056 | 0.1368 |
|  |  | P value summary | ns | ns |
| PRNT<sub>90</sub> SARS2 | Spearman r | r | 0.7922 | 0.8525 |
|  | P value | P (two-tailed) | 0.0004 | <0.0001 |
|  |  | P value summary | *** | **** |

ns, non-significant $P > 0.05$ ; \* reflects significance level: \*\*, $P \leq 0.01$ ; \*\*\*, $P \leq 0.001$ ; \*\*\*\*, $P \leq 0.0001$

## Discussion

Validated SARS-CoV-2 serological assays are currently lacking, yet urgently needed for contact tracing, epidemiological and vaccine evaluation studies. Since the N and the S proteins are the main immunogenic CoV proteins, we developed ELISA-based assays, which were able to detect antibodies to these two proteins along with the two spike domains, S1^A^ and RBD. Those correlated strongly with virus neutralizing antibodies detected by a PRNT_50_ assay. Since the majority of the human population has antibodies against the four endemic human coronaviruses, it was crucial to verify the specificity of these assays to avoid false positive results. Additionally, the two zoonotic coronaviruses, SARS-CoV and MERS-CoV, are also beta-coronaviruses, raising potential for cross-reactivity. Among the spike protein antigens tested, the S1 was more specific than S in detecting SARS-CoV-2 antibodies, as MERS-CoV-S cross-reactive antibodies were detected in the serum of one of the COVID-19 patients which was not seen when MERS-CoV S1 was used for testing. This could be explained by the high degree of conservation in the CoV S2 subunit relative to the S1 (**Table 2**). Therefore, consistent with our earlier findings for MERS-CoV serology (6), S1 is a specific antigen for SARS-CoV-2 diagnostics.

When testing the specificity S1 or its RBD for detecting SARS-CoV-2 antibodies, none of the sera form the validation cohorts (A-E) showed any reactivity; except for SARS-CoV patients sera. This - not-unexpected - cross-reactivity resulted from the high degree of similarity between the S1 and RBD of the SARS-CoV and SARS-CoV-2 (**Table 2**). However, SARS-CoV has not circulated in the human population since 2003, i.e. 17 years ago, and an earlier study reported waning of SARS-CoV-specific antibodies which made them undetectable in serum samples of 91% (21/23) of samples tested 6 years following infection (11). It is therefore unlikely that antibodies to this virus are present in the population and thus there could hardly be a chance that false positives result from SARS-CoV-antibodies reactivity. Meanwhile, we made use of the high degree of similarity between the SARS-CoV and SARS-CoV-2 proteins for the development of our inhouse N ELISA, where we used SARS-CoV N (90% similar to SARS-CoV-2) as an antigen. The N-ELISA could detect SARS-CoV-2-specific antibodies with high specificity and sensitivity. Using the three different validated ELISAs, we found that antibody levels were higher following the severe infection compared to the mild ones; similar findings has been reported earlier for MERS-CoV (12, 13). However, this needs to be confirmed with a larger cohort of patients with varying degrees of severity, while it still highlights the potential need of a sensitive assay to avoid missing those with milder infections in epidemiological studies. Among the 3 inhouse ELISAs tested, the RBD and N ELISAs were more sensitive than S1 ELISA in detecting antibodies in mildly infected patients and showed stronger correlation with PRNT_50_ titers. Therefore, detecting antibodies against two different antigens might be needed to confirm the findings and avoid false negatives in surveillance studies. However, the sensitivities of the assays need to be further validated with a larger cohort.

We further validated beta-versions of an IgA and an IgG S1 commercial ELISA in two different labs. While the IgA-based ELISA showed higher sensitivity than the IgG-based ELISA, the opposite was true for the specificity where the IgG ELISA was more specific than the IgA ELISA. Yet, we noted some cross reactivity in both ELISAs with serum samples from the same two HCoV-OC43 patients that cross reacted in a MERS-CoV S1 IgG ELISA (6) despite the different antigen coated. This indicates a response to another protein which could be in the blocking or coating matrix, apart from the specific antigen coated, resulting in this consistent false positive result.

Overall, the assays developed and validated in this study could be instrumental for patient contact tracing, serosurveillance studies, as well as vaccine evaluation studies. However, since various studies will be carried out in different labs, it is crucial to calibrate and standardize assays developed by different labs using well defined standard references as a part of diagnostic assay validation. This is not only needed to reduce interassay variability, but to also harmonize the results obtained from different labs using various assays (14). This is crucial for better comparison and interpertaion of results from different studies as well as evaluation of vaccine trials, allowing for uniform assessment of immunogenicity, efficacy and better understanding of correlates of immune protection (15). Thus, setting up reference panels is a vital element in our preparedness approaches to emerging viruses.

## Data Availability

All data referred to in the manuscript are available form corresponding authors upon reasonable request.

## Acknowledgments

This work was supported by the Zoonoses Anticipation and Preparedness Initiative (ZAPI project; IMI grant agreement no. 115760), with the assistance and financial support of IMI and the European Commission, in-kind contributions from EFPIA partners.

